# Deep Learning vs manual techniques for assessing left ventricular ejection fraction in 2D echocardiography: validation against CMR

**DOI:** 10.1101/2022.07.26.22278059

**Authors:** Eric Saloux, Alexandre Popoff, Hélène Langet, Paolo Piro, Camille Ropert, Romane Gauriau, Romain Stettler, Mihaela Silvia Amzulescu, Guillaume Pizaine, Pascal Allain, Olivier Bernard, Amir Hodzic, Alain Manrique, Mathieu De Craene, Bernhard L. Gerber

## Abstract

**Objective:** To evaluate accuracy and reproducibility of 2D echocardiography (2DE) left ventricular (LV) volumes and ejection fraction (LVEF) estimates by Deep Learning (DL) vs. manual contouring and against CMR.

**Background:** 2DE LV manual segmentation for LV volumes and LVEF calculation is time consuming and operator dependent.

**Methods:** A DL-based convolutional network (DL1) was trained on 2DE data from centre A, then evaluated on 171 subjects with a wide range of cardiac conditions (49 healthy) – 31 subjects from centre A (18%) and 140 subjects from centre B (82%) – who underwent 2DE and CMR on the same day. Two senior (A_1_ and B_1_) and one junior (A_2_) cardiologists manually contoured 2DE end-diastolic (ED) and end-systolic (ES) endocardial borders in the cycle and frames of their choice. Selected frames were automatically segmented by DL1 and two DL algorithms from the literature (DL2 and DL3), applied without adaptation to verify their generalizability to unseen data. Interobserver variability of DL was compared to manual contouring. All ESV, EDV and EF values were compared to CMR as reference.

**Results:** 50% of 2DE images were of good quality. Interobserver agreement was better by DL1 and DL2 than by manual contouring for EF (Lin’s concordance = 0.9 and 0.91 vs. 0.84), EDV (0.98 and 0.99 vs. 0.82), and ESV (0.99 and 0.99 vs. 0.89). LVEF bias was similar or reduced using DL1 (-0.1) vs. manual contouring (3.0), and worse for DL2 and DL3. Agreement between 2DE and CMR LVEF was similar or higher for DL1 vs. manual contouring (Cohen’s kappa = 0.65 vs. 0.61) and degraded for DL2 and DL3 (0.48 and 0.29).

**Conclusion:** DL contouring yielded accurate EF measurements and generalized well to unseen data, while reducing interobserver variability. This suggests that DL contouring may improve accuracy and reproducibility of 2DE LVEF in routine practice.

## Introduction

Accurate and reproducible echocardiographic assessment of left ventricular (LV) volumes and ejection fraction (EF) is crucial in clinical decision-making and risk stratification (1–6). Hence LVEF thresholds are used for decision making in heart failure (7) and coronary artery (8) and valvular heart (9) diseases. Simpson’s method from two-dimensional (2D) 2- and 4-chamber views is currently the preferred approach for evaluation of LV volumes and EF by echocardiography. Yet it is time consuming, subject to wide interobserver variability (10,11), and the reproducibility is highly affected by various factors such as operator experience and image quality. Accordingly, 2D echocardiography has shown to be less reproducible than cardiac magnetic resonance (CMR)(12), which is currently considered the reference standard for evaluation of LVEF and volumes. This approach however suffers from higher costs and less frequent availability.

Deep learning (DL) allows automated contour detection offering the promise of faster and potentially more accurate and reproducible evaluation of LV volumes and EF by echocardiography (3,7,13,14). In this work, we developed a new DL algorithm for manual contouring based on a U-Net convolutional network architecture, using an anonymised database of echocardiographic images. The aim of the present study was to evaluate the generalizability and accuracy of this new algorithm relative to manual contouring and against cardiac MR volumes and EF as a reference. We evaluated our algorithm using a set of multimodal data from 171 subjects from two centres. We also compared the automated contouring and resulting LV volumes to their manual counterparts, as obtained by different junior and senior observers across the two centres and used CMR as an independent 3D modality to evaluate differences in bias between manual and automated contouring. Finally, we benchmarked our DL algorithm against two other DL implementations from the literature and made the 2DE database, CMR LVEF values and the Simpson bi-plane code publicly available for reproducibility of this paper and further benchmarks.

## Methods

### Study Design

The present study is a retrospective analysis of echocardiography and CMR data from participants previously enrolled in prospective trials performed either at Centre Hospitalier Universitaire de Caen Normandie (denoted as clinical centre A) or at Cliniques Universitaires Saint-Luc (denoted as clinical centre B), following approval by an ethic committee. The studies were approved by the IRB in charge (either Comité de Protection des Personnes Nord-Ouest III, Caen, France or Comité Ethique Hospitalo-Facultaire de l’Université Catholique de Louvain, Brussels, Belgium). The retrospective use of the data satisfies the European Union (EU) General Data Protection Regulation (GDPR) requirements. Data from participants who had undergone 2D echocardiography and cardiac CMR within 24 hours and who were found in sinus rhythm were analysed in the present study, resulting in a database of 171 subjects in total. Among those, there were 49 healthy volunteers and 122 patients with various cardiac pathologies: 37 with an ischaemic heart disease and a previous myocardial infarct, 35 with a non-ischaemic dilated cardiomyopathy, 39 with a valvular heart disease (including 19 aortic stenoses, 17 mitral regurgitations, 2 mitral repairs and 1 aortic regurgitation), and 11 with a hypertrophic cardiomyopathy. There was no prior selection based on image quality, thus reflecting a realistic range of echogenicity and artefacts in the database. For each subject, demographic and anthropometric data (age, sex, height, weight), diastolic and systolic blood pressures, and cardiovascular risk factors were also extracted.

### 2D transthoracic echocardiography

Standardized comprehensive transthoracic echocardiographic examinations had been acquired according to established guidelines (15) using Philips IE 33 and EPIQ 7 ultrasound system equipped with a X5-1 transducer in harmonic imaging (Philips Medical Systems, Andover, MA, United States), and stored on a PACS server (Intellispace Cardiovasular (ISCV), Philips Medical Systems, Andover, MA, United States).

### Echocardiography measurements

#### Manual measurements

Echocardiographic images were anonymized, exported in DICOM format and analysed off-line. Three successive cardiac cycles were available for apical 4- and 2-chamber views. Two senior (A_1_: ES) and (B_1_: BLG) and one junior (A_2_: RS) cardiologists manually contoured the ED and ES endocardial borders in the cardiac cycle and on the image frames of their choice, while blinded to quantitative outcomes. Senior cardiologists had more than 20 years of experience in echocardiography, the junior cardiologist had 3 months of training. For DL analysis, a deep convolutional neural network (U-Net) model was used to segment the LV cavity on the frames selected by the three observers. The same biplane Simpson’s method was used to compute all LV volumes and EF (see *Volumes computation* below). The study protocol is summarized in (Fig 1). Data were analysed in two independent ways (manual EF, and DL-EF for the 3 compared DL algorithms) for all frames selected by the 3 observers. Observers were blinded to other manual and automated quantifications. Manual analysis was performed with a custom Python script running a Web browser-based interface to present images in random order and to save the contours. The observers first selected a cycle among the three consecutive cycles, then determined ED and ES within this cycle. An interactive graphical interface allowed the contouring of the cavity on both the AP2 and AP4 views. All observers were instructed to respect the following conventions: i) include trabeculae and papillary muscles in the LV cavity; ii) keep consistency in excluding/including tissue between ED and ES; iii) terminate the contours in the mitral valve plane on the ventricular side of the bright ridge, at the points where the valve leaflets are hinging. Automated segmentation was performed using three deep learning algorithms (see below). The algorithms were run on all frames selected by observers. Therefore, DL measurements will be presented systematically for the 3 observers (junior A_1_ and senior A_2_ and B_1_).

**Fig 1.**
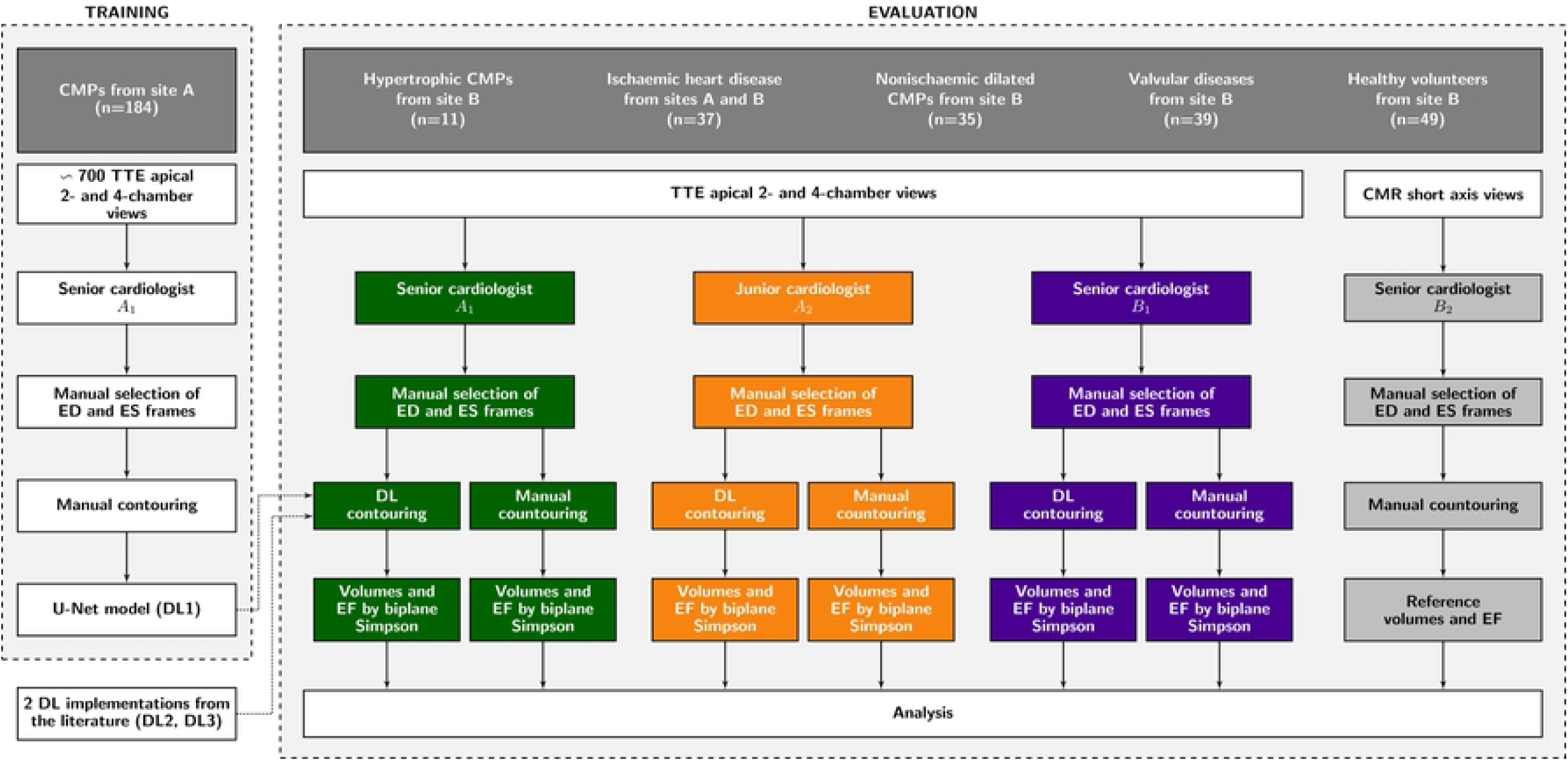
Flowchart of the study. The deep convolutional neural network (U-Net) model was trained on about 700 apical 2- and 4-chamber views from site A. The performance of Deep learning (DL) analysis using the U-Net model was then evaluated on data from 171 subjects from sites A and B, who underwent both transthoracic echocardiography and cardiac magnetic resonance (CMR). CMP = cardiomyopathies. DL = Deep Learning. ED = end-diastole. ES = end-systole.

#### Volumes computation

A standard Simpson’s rule for biplane EF computation was implemented according to (16) and applied to manual and DL segmentations. The code for this Simpson’s implementation is made publicly available on a git repository. The first step is first to find for both the AP2 and AP4 images a rotated bounding box fully encompassing the input contour. The second step is to find the apex and mitral points. The apex is defined as the contour point the closest to the middle of the bounding box top edge. The mitral point is found as the contour point that intersects the long axis direction of the bounding box. Mitral and apex points define the long axis segment in both AP2 and AP4 images. These long axis segments are divided into twenty points at which two radiuses are cast towards both sides of the input contour. By summing the N ellipsoidal cylinders from the AP2 and AP4 contours, one obtains the EDV and ESV values.

#### Image quality assessment

To assess the feasibility of DL with respect to image quality, senior observer B_1_ ranked the image quality of all frames (i.e., both ES and ED frames for AP2 and AP4) into three categories: good, meaning the endocardial wall was visible for the whole LV; fair, meaning the endocardium had to be visually interpolated at some locations; and poor, meaning that the frame was not of sufficient quality for being quantified. Senior observer A_1_ also classified all DL segmentations according to whether he would have edited or not the automated contouring. Finally, A_1_ also counted the cases for which DL outperformed Junior Observer A_2,_ assessing if DL was worse, as good or better than A_2_’s contouring for quantification purposes.

### Deep learning algorithms

A U-Net architecture was trained to segment the LV cavity mask using ED and ES manual contouring from senior cardiologist A_1_ on an independent database of about 700 anonymised apical AP2 and AP4 views from 237 subjects. That database consisted of patients with ischemic cardiomyopathy (59%), dilated cardiomyopathy (6%), valvular pathologies (9%), hypertrophic cardiomyopathy (3%). Remaining subjects in the training database underwent 2D echo because of arterial hypertension or other cardiovascular risk factors. For all included subjects, image quality had been deemed satisfactory by A_1_ to be manually contoured in the AP2 and AP4 views. The convolutional neural network architecture followed the standard U-Net pattern, stacking elementary blocks of convolutional, activation and batch normalization layers (17). The network took as input the full scan-converted B-Mode image, resized to a 192×256 size. The activation of the final layer (sigmoid) outputted a continuous mask image that took values between 0 and 1. After thresholding this result at 0.5, the largest contour was extracted for the AP2 and AP4 ES and ED frames. This method is referred to as DL1 in the remainder of this paper.

For benchmarking DL1 against other techniques, two other DL implementations from the literature were evaluated. First, the e-Net architecture from Leclerc et al. (13) was applied on the same frames as the DL method. This method is referred to as DL2 in the remainder of this paper. The DL2 network was trained on GE images, on the CAMUS public database (13). The weights of the network were used as such, without any adaptation. Finally, the method of Zhang et al. (18) was applied similarly without any retraining. This model is referred to as DL3.

### Magnetic resonance imaging

All subjects had undergone a standardized CMR myocardial function study on a 3T scanner (Achieva, Philips Medical Systems, Best, the Netherlands). The CMR exam was performed within a 24 hours window from the echocardiographic exam. 10-12 consecutive short axis images covering the entire LV, and respectively one 2- 3- and 4 chambers long-axis cine SSFP images were acquired for assessment of myocardial function. CMR RV and LV volumes and EF were computed using Segment version 2.2 (*http://segment.heiberg.se*) (19) or Medis software (Medical Imaging Systems, Leiden, the Netherlands) from short-axis cine images by semi-automatically contouring the endo- and epicardial contours in the end-diastolic (ED) and end-systolic (ES) phases. These quantifications were performed by an independent EuroCMR level III certified operator (MSA) blinded to the quantitative findings of echocardiographic operators. Papillary muscles and trabeculations were included as blood volume in the cavity contour.

### Statistical analysis

Statistical analyses were performed using the epiR and psych R packages. Continuous variables are presented as mean values±SD, categorical variables as counts and percentages. Continuous variables were compared using the independent sample Student t test if normally distributed, or else using either the Wilcoxon signed rank test (paired data) or the Mann-Whitney (unpaired data) tests. A p-value p<0.05 was considered statistically significant. Agreements between 2D-echo DL and manual segmentations for each observer was assessed using the DICE similarity coefficient computed as 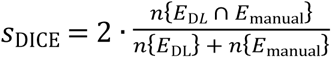 where *E*_DL_ and *E*_manual_ are the sets of pixels found within the LV cavity by DL and manual contouring, and *n*{⋅} is the number of pixels in a given set. DICE is a standard measure to compare the overlap of two binary segmentation masks. Both interobserver and echo vs. CMR agreements were measured with Lin’s concordance correlation coefficient (CCC) for EDV, ESV, and EF. To better evaluate the impact of different EF values using either DL vs. manual contouring or CMR vs. 2D echo, we categorized EF into three thresholds: EF<40%, 40%≤EF≤50%, EF>50% matching the guidelines for the LVEF stratification of HF patients (20). We then measured agreement to classify subjects into these 3 groups among each observer and DL by 2D echo and CMR using Cohen’s kappa (κ) coefficient.

## Results

### Study Population

Table 1 presents baseline characteristics of the study population. The validation cohort of this study was composed of n=31 (18%) patients from centre A and n=140 (82%) patients from centre B. The population had a wide range of LV ejection fraction and volumes. There were no significant differences in hemodynamics between echo and CMR studies. As expected, there were significant differences in age, EDV, ESV and EF among subjects with different cardiac conditions. Image quality of echo images was rated by B_1_ as good (resp. fair and poor) for 50% (resp. 42% and 8%) of the frames composing the dataset.

**Table 1.**
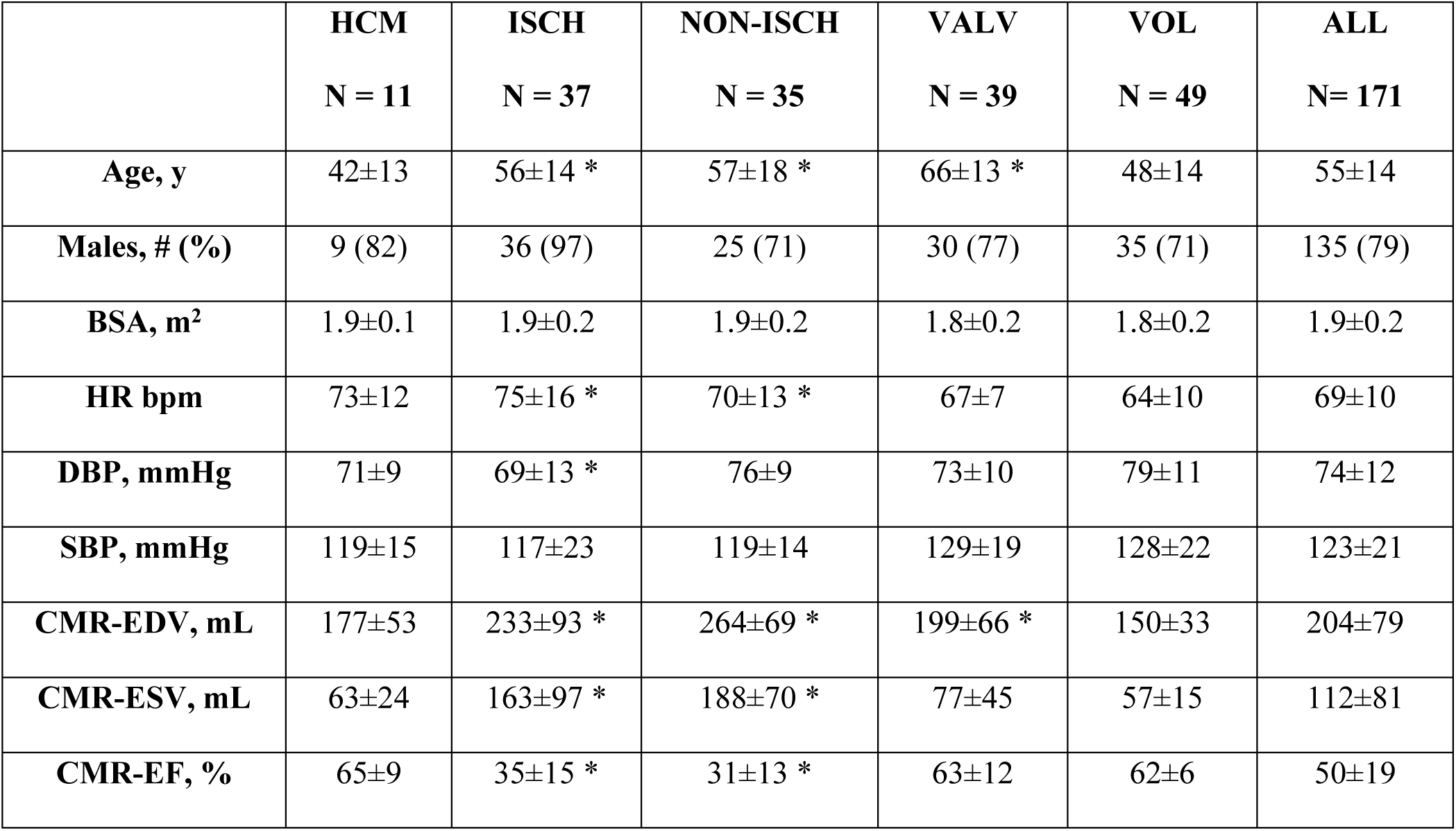
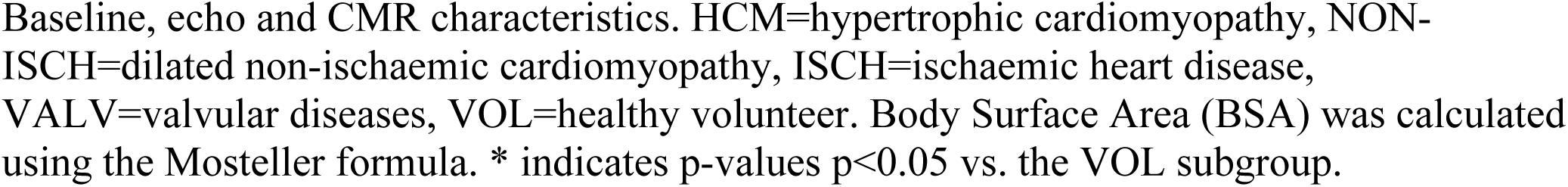
Patient population characteristics.

|  | **HCM**<br>**N = 11** | **ISCH**<br>**N = 37** | **NON-ISCH**<br>**N = 35** | **VALV**<br>**N = 39** | **VOL**<br>**N = 49** | **ALL**<br>**N = 171** |
| --- | --- | --- | --- | --- | --- | --- |
| **Age, y** | 42±13 | 56±14 * | 57±18 * | 66±13 * | 48±14 | 55±14 |
| **Males, # (%)** | 9 (82) | 36 (97) | 25 (71) | 30 (77) | 35 (71) | 135 (79) |
| **BSA, m<sup>2</sup>** | 1.9±0.1 | 1.9±0.2 | 1.9±0.2 | 1.8±0.2 | 1.8±0.2 | 1.9±0.2 |
| **HR bpm** | 73±12 | 75±16 * | 70±13 * | 67±7 | 64±10 | 69±10 |
| **DBP, mmHg** | 71±9 | 69±13 * | 76±9 | 73±10 | 79±11 | 74±12 |
| **SBP, mmHg** | 119±15 | 117±23 | 119±14 | 129±19 | 128±22 | 123±21 |
| **CMR-EDV, mL** | 177±53 | 233±93 * | 264±69 * | 199±66 * | 150±33 | 204±79 |
| **CMR-ESV, mL** | 63±24 | 163±97 * | 188±70 * | 77±45 | 57±15 | 112±81 |
| **CMR-EF, %** | 65±9 | 35±15 * | 31±13 * | 63±12 | 62±6 | 50±19 |

Baseline, echo and CMR characteristics. HCM=hypertrophic cardiomyopathy, NON-ISCH=dilated non-ischaemic cardiomyopathy, ISCH=ischemic heart disease, VALV=valvular diseases, VOL=healthy volunteer. Body Surface Area (BSA) was calculated using the Mosteller formula. \* indicates p-values $p < 0.05$ vs. the VOL subgroup.

### Feasibility of DL-based contouring

DL computation using the DL1 network of EDV and ESV and EF was feasible in all subjects and images and took about 60 ms per image (including biplane Simpson’s computation). Typical examples of DL1 contouring are shown in (Fig 2). Reviewer A_1_ considered DL1 segmentations as acceptable, and not requiring further editing, in 70% of the frames (64% for fair/poor images). A_1_ also considered 16.4% of the frames to be better segmented by DL1 than manual segmentation by the junior observer A_2_. For the latter, A_1_ was not blinded to which method was used to produce the segmentation result. In only 6 cases (3%) DL segmentation was considered to have failed as illustrated in (S1 Fig). In a patient with non-ischemic dilated CMP and a patient with mitral regurgitation, a hyper-intense valve apparatus in the image disrupted the DL1 segmentation and yielded a cavity segmentation with a highly irregular shape. In two patients (with aortic stenosis and mitral regurgitation), a partly non-visible endocardial wall impeded automatic segmentation. In the two last outliers, one hypertrophic and aortic stenosis, with poor LV function and low EF, local DL1 segmentation errors had a higher impact than in subjects with good LV function.

**Fig 2.**
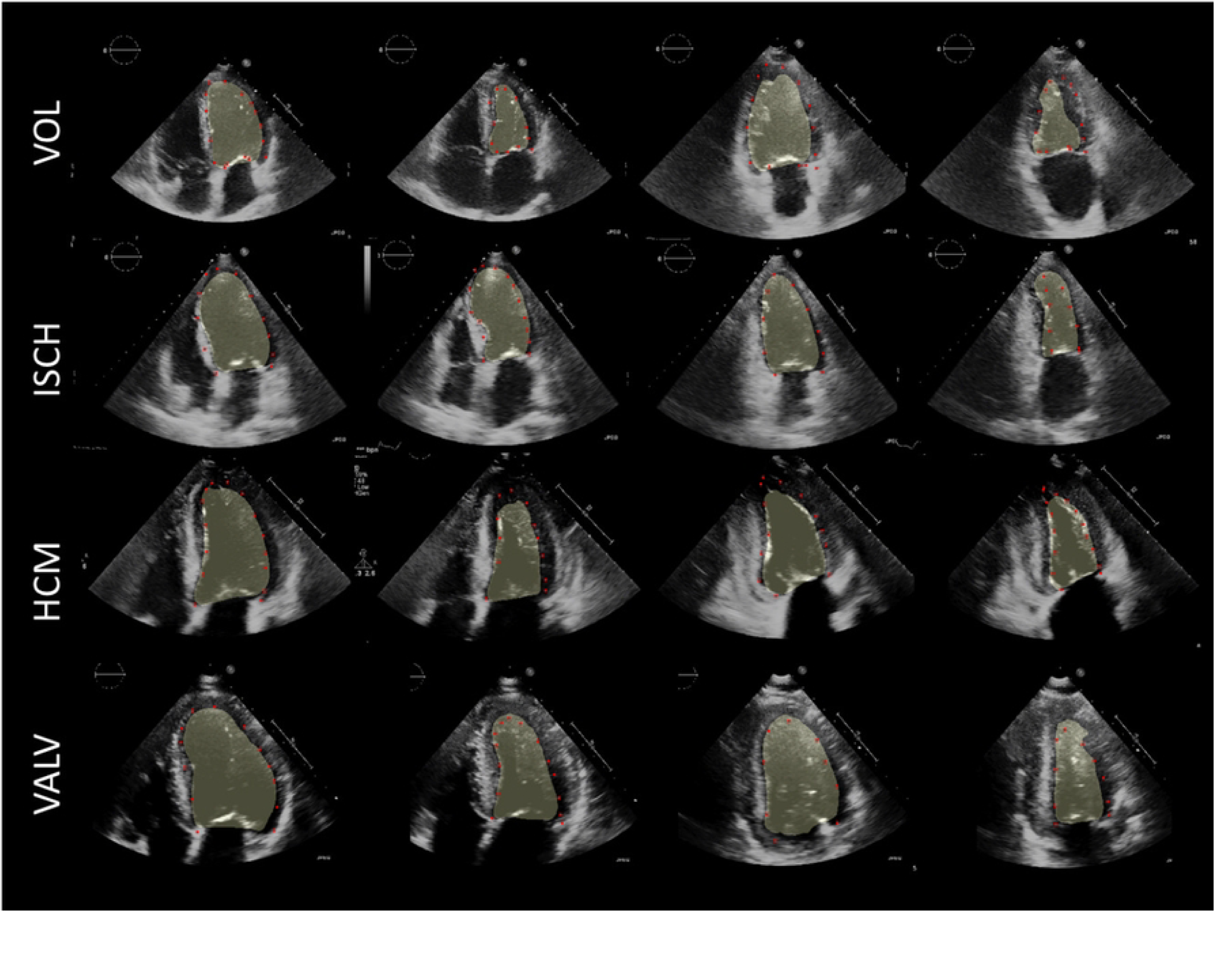
Overlay of typical DL1 segmentations (yellow mask) and manual contouring by senior observer B_1_ (red dotted contour) for four representative subjects. From top to bottom: healthy volunteer (VOL), subject with a ischaemic heart disease (ISCH), subject with a hypertrophic cardiomyopathy (HCM), and subject with an aortic stenosis (VALV). From left to right: end-diastolic frame in apical 4-chamber view, end-systolic frame in apical 4-chamber view, end-diastolic frame in apical 2-chamber view, and end-systolic frame in apical 2-chamber view.

### Agreement of DL and manual contouring

(Fig 3) shows the DICE values spread when comparing DL1 vs. manual contours on the (ES, ED) × (AP2, AP4) frames of every observer. For all echo views and observers, the average DICE values were over 90%. However, a more elevated spread appeared in the junior (A_2_) compared to the two senior (A_1_, B_1_) observers. In addition, the AP4 view showed a higher consistency between manual and DL results. Similarly, within each echo view, lower DICE values were found in ES than in ED (with the exception of observer B_1_ in the A2C view).

**Fig 3.**
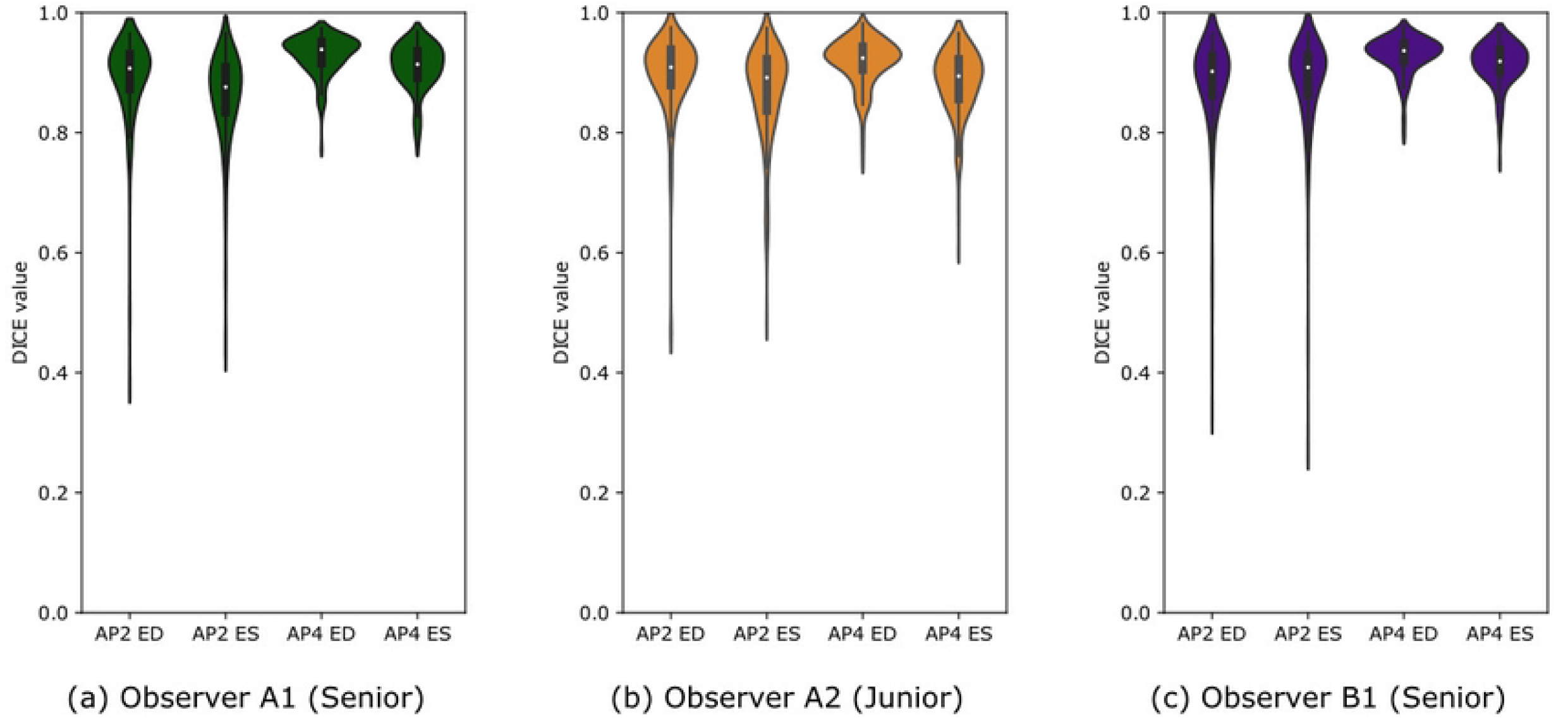
DL1 vs. manual comparison. Left-ventricular cavity overlap as measured by the DICE similarity coefficient between manual and DL1 contouring in apical 2- and 4-chamber views for all observers.

### Comparison of EF, ESV and EDV values from DL-based contours and manual contours

Table 2 lists the computed ranges of EF, EDV and ESV values from echo images for the whole population and disaggregated by healthy and pathology groups, when quantified either by the senior and junior observers or DL1. EF values for most pathological groups were found non significantly different when measured by A_1_ or DL1. DL1 values of EF were also, but to a lesser extent, consistent with A_2_ and B_1_, who both reported higher overall EF values. For EDV and ESV, there were significant differences between observers, with junior observer A_2_ providing systematically lower EDV and ESV estimates than senior observers A_1_ and B_1_.

### Interobserver variability

Interobserver agreement (for EF, ESV and EDV) was significantly better for DL1 than manual contouring. As illustrated in (Fig 4), this effect was most dominant for LV-EDV where there was particularly poor senior-junior (compared to senior-senior) agreement of EDV values**Error! Reference source not found**. S1 Table quantifies inter-observer agreement in manual vs. DL for the 3 DL algorithms. It shows both DL1 and DL2 reached excellent inter-observer agreement (Lin’s CCC >0.9 for EDV, ESV and EF) that compared favourably to manual inter-observer agreement (Lin’s CCC < 0.9 for EDV, ESV and EF)

**Fig 4.**
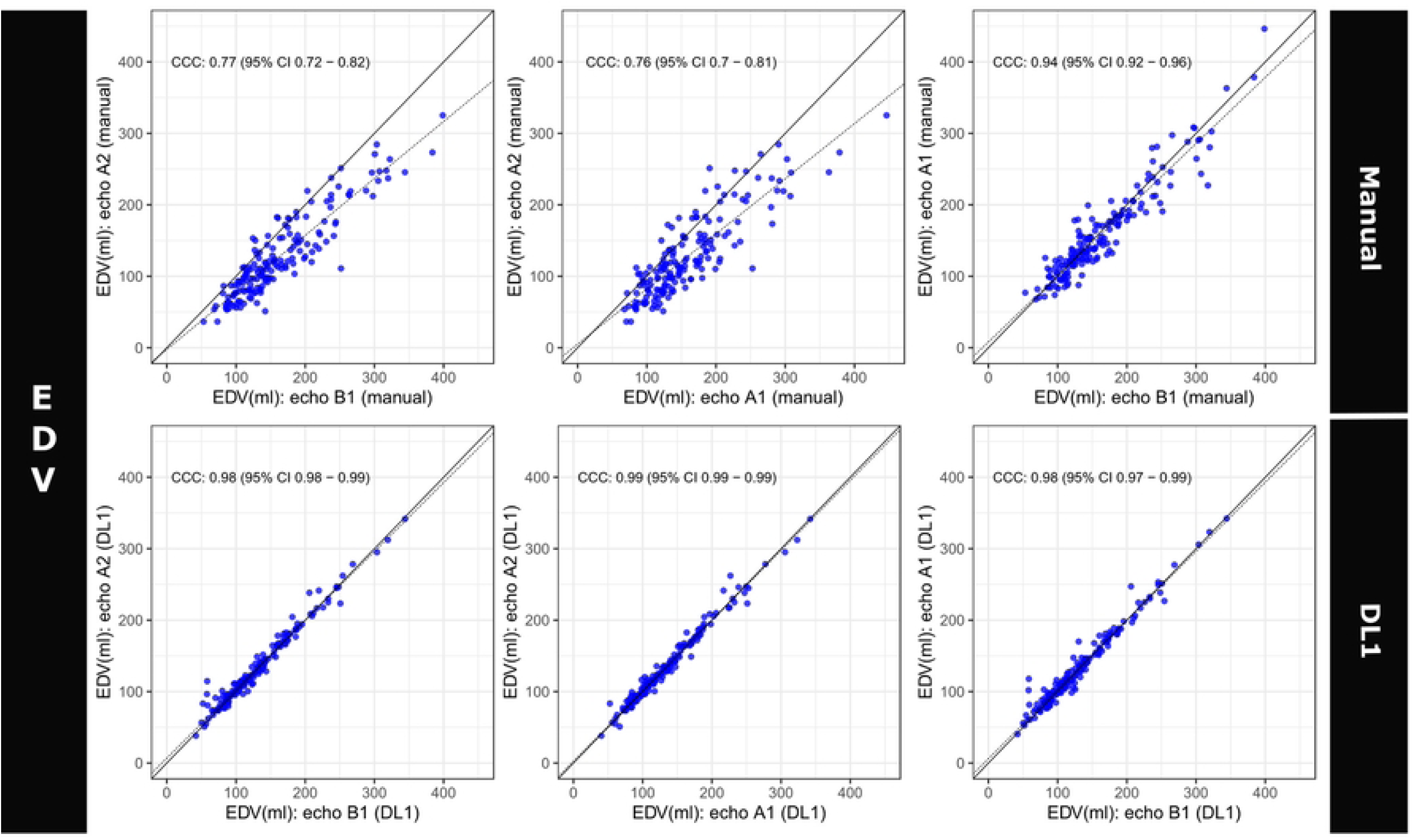
Lin’s concordance correlation plots between observers (top) and for DL1 (bottom, on the frames quantified by all observers) for EDV in 2D echo.

### Agreement of Manual and DL1 measurements with CMR

As shown in Table 2, EF values were consistent between CMR and echo for all observers using either manual contouring or DL1. Correlation and Bland-Altman plots for EF are shown in (Fig 5). Lin’s CCC between echo-EF and CMR-EF improved for A_2_ using DL but degraded for senior observers A_1_ and B_1_. The EF bias was reduced with DL1 compared to manual contouring for junior observer A_2_ (-0.1±10.0 vs. 3.7±9.6) and senior observer B_1_ (-0.7±13.1 vs 5.9±8.1) but remained unchanged for A_1_ (0.5±11.3 vs -0.5±8.6). Regarding volume measurements, (Fig 6) shows that all echo-based ESV and EDV (using either manual or DL1 for all observers) values were underestimated w.r.t. CMR. Correlation between EDV by senior observers and CMR were good (Fig 6) with CCC values over 0.7, higher than the junior cardiologist A_2_ (0.45). This value slightly improved for A_2_ using DL (0.51). In comparison, DL1 ESV values were in better agreement with CMR (CCC > 0.7).

**Table 2.:** LV volumes and EF by all observers and modalities.

|  | HCM | ISCH | NON-ISCH | VALV | VOL | ALL |
| --- | --- | --- | --- | --- | --- | --- |
| **End-diastolic volumes [mL]** |  |  |  |  |  |  |
| **Senior A1** | 120±39 | 189±69 * | 203±64 * | 155±58 * | 117±24 | 159±64 ‡ |
| **Senior B1** | 108±33 | 196±70 * | 204±65 * | 162±61 * | 122±24 | 163±65 † |
| **Junior A2** | 72±24 *†‡ | 157±59 *†‡ | 154±56 *†‡ | 128±61 †‡ | 100±30 †‡ | 128±57 †‡ |
| **DL** | 91±27 † | 165±57 *†‡ | 165±56 *†‡ | 122±43 †‡ | 104±23 †‡ | 133±53 †‡ |
| **CMR** | 177±53 †‡ | 233±93 *†‡ | 264±69 *†‡ | 199±66 *†‡ | 150±33 †‡ | 204±79 †‡ |
| **End-systolic volumes [mL]** |  |  |  |  |  |  |
| **Senior A1** | 52±23 ‡ | 120±65 *‡ | 137±57 *‡ | 69±41 *‡ | 47±14 ‡ | 87±58 ‡ |
| **Senior B1** | 35±17 † | 113±67 *† | 130±63 *† | 59±32 † | 41±12 † | 78±59 † |
| **Junior A2** | 25±12 †‡ | 92±56 *†‡ | 96±46 *†‡ | 50±29 †‡ | 38±15 † | 64±46 †‡ |
| **DL** | 39±19 † | 105±63 *†‡ | 113±52 *†‡ | 53±23 *† | 38±12 †‡ | 71±52 †‡ |
| **CMR** | 63±24 ‡ | 163±97 *†‡ | 188±70 *†‡ | 77±45 †‡ | 57±15 †‡ | 112±81 †‡ |
| **Ejection fractions [%]** |  |  |  |  |  |  |
| **Senior A1** | 58±12 ‡ | 40±14 *‡ | 34±12 * | 58±13 ‡ | 61±6 ‡ | 50±16 ‡ |
| **Senior B1** | 69±10 † | 46±16 *† | 38±14 * | 64±11 † | 67±6 † | 56±17 † |
| **Junior A2** | 66±9 † | 45±16 *† | 40±11 * | 62±10 | 62±7 ‡ | 54±15 †‡ |
| **DL** | 57±14 | 41±17 *‡ | 33±15 *‡ | 56±19 *‡ | 64±7 †‡ | 50±19 †‡ |
| **CMR** | 65±9 | 36±16 *†‡ | 31±13 *‡ | 63±12 †‡ | 62±6 | 50±19 ‡ |

Left ventricle volumes and ejection fraction in the different groups of cardiac conditions as measured by manual contouring and DL contouring from 2D echocardiography and semi-automated contouring from CMR. See group definitions in **Error! Reference source not found.** \* indicates p<0.05 vs. the VOL subgroup. † indicates p-values p<0.05 vs. A<sub>1</sub>. ‡ indicates p-values p<0.01 vs. B<sub>1</sub>.

**Fig 5.**
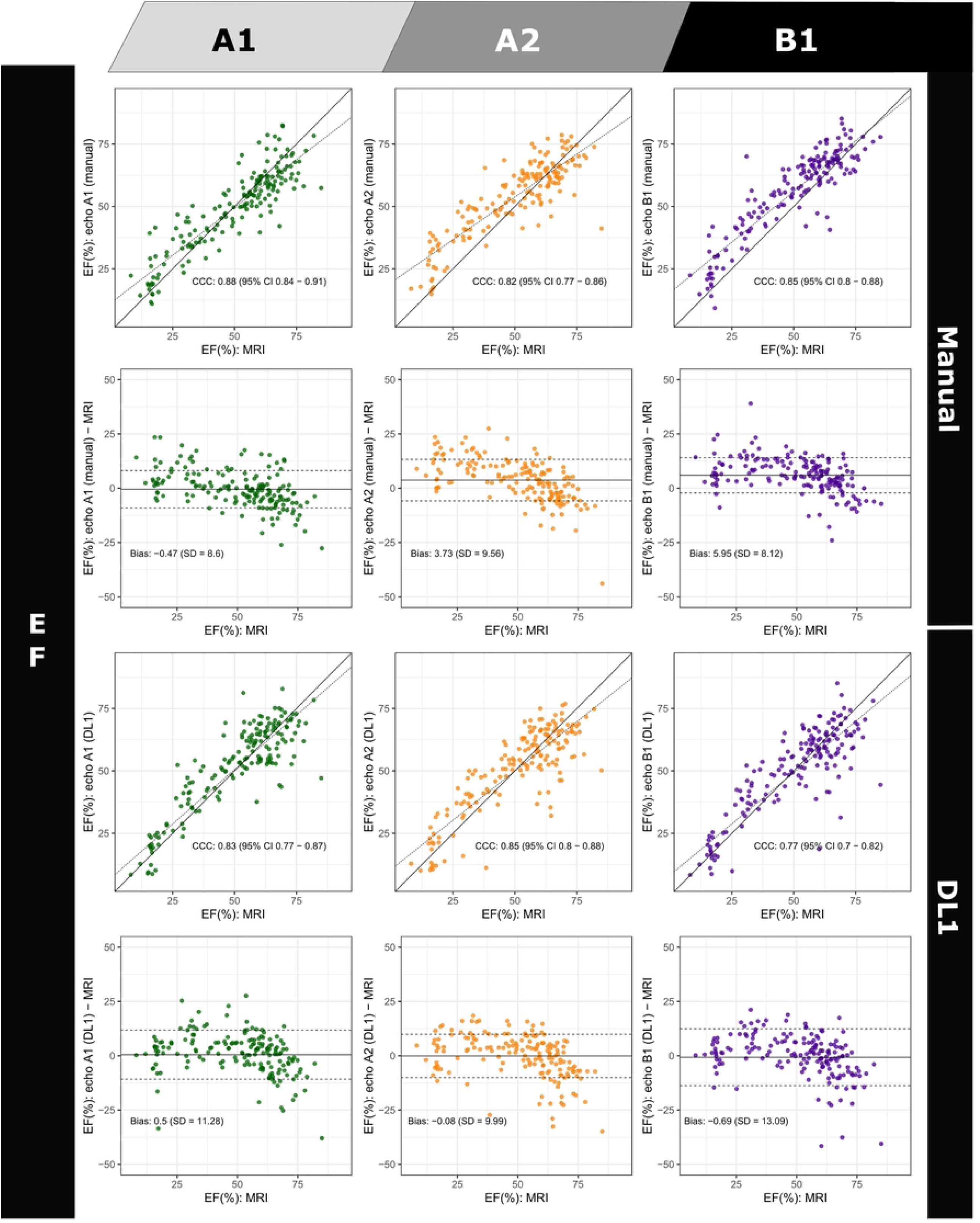
Lin’s concordance correlation coefficient and Bland-Altman plot for EF comparing manual and DL1 estimates for each observer in 2D echo to MRI.

**Fig 6.**
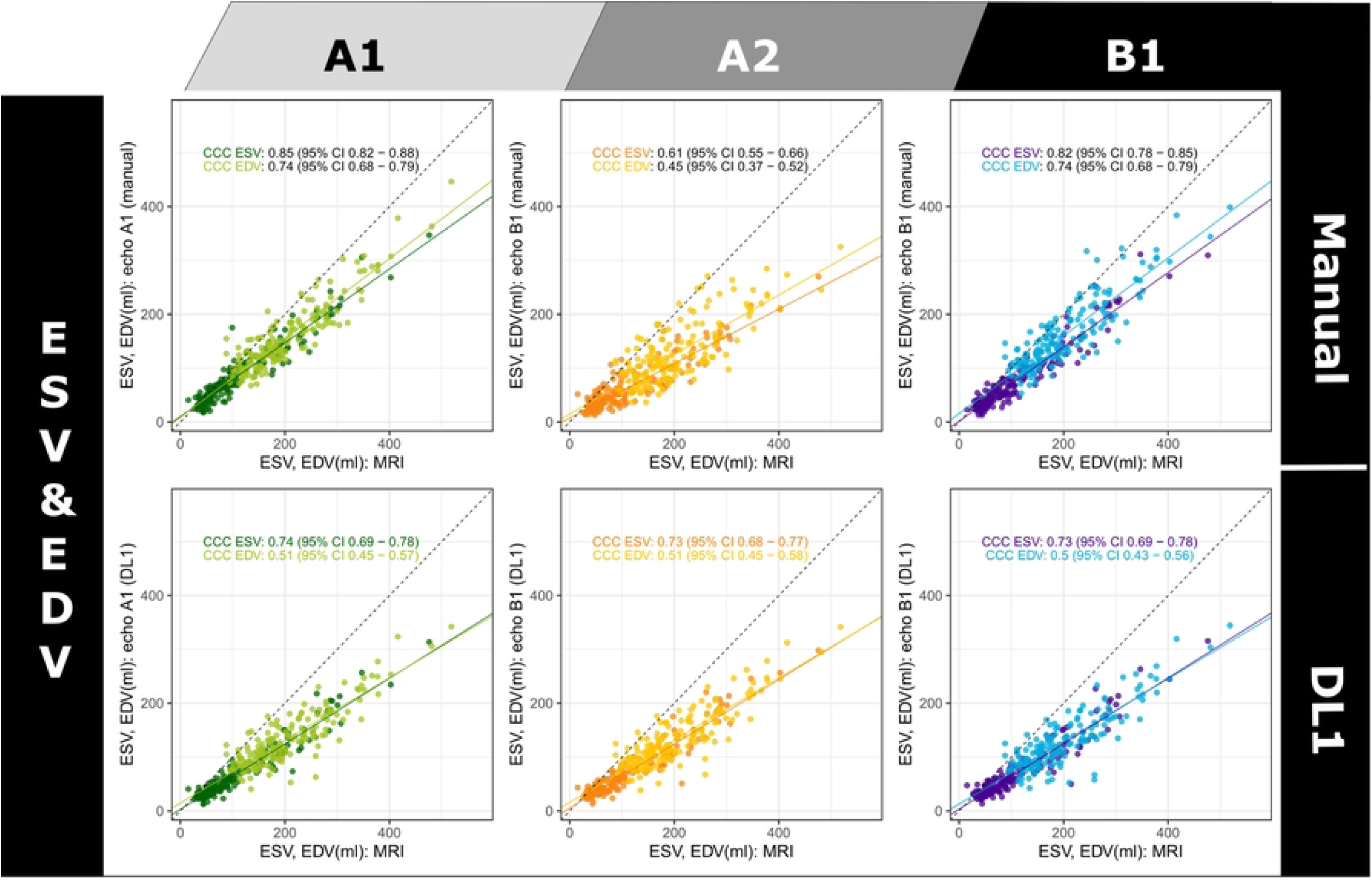
Lin’s concordance correlation coefficient plots for LV EDV and LV ESV between CMR and 2D echo for all observers and DL1 applied to the frames of each observer. (A_1_ [senior], A_2_ [junior] and B_1_ [senior]).

### Comparison to other DL methods

(Fig 7) compares DL1, DL2 and DL3 agreement measurements on all frames selected by observers with CMR. DL1 and DL2 exhibited similar Lin’s CCC for volume evaluation, while DL3 performed less well (0.51 resp. 0.49 vs. 0.22 for EDV, and 0.73 resp. 0.74 vs. 0.53 for ESV). EF values were not valid in 12.7% of the cases for DL3 vs. 0.78% for DL1 and 1.56% for DL2. All three DL methods showed acceptable Pearson correlation (0.86 for DL1, 0.72 for DL2, and 0.57 for DL3), but only DL1 and DL2 showed acceptable agreement with CMR. DL1 achieved higher agreement than DL2 due to a reduced bias and SD, thus contrasting with their more homogeneous EDV / ESV findings.

**Fig 7.**
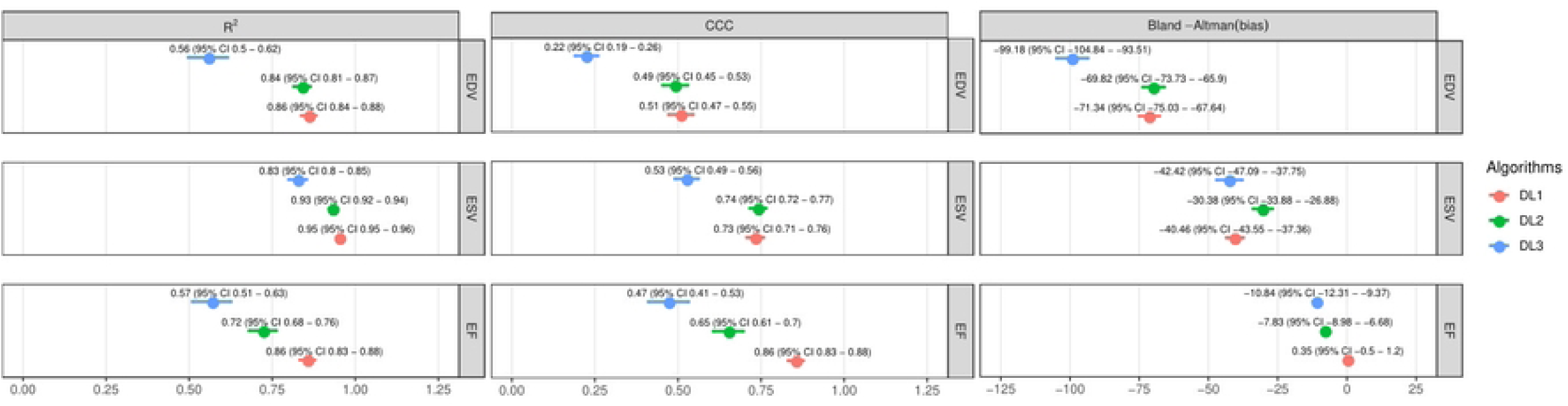
Agreement between CMR and 2D echo for all observers and the three DL techniques applied to the frames of each observer. (A1 [senior], A2 [junior] and B1 [senior]). DL1 is the network proposed in this paper, DL2 is the e-Net architecture from (13), DL3 is the network proposed in (18).

### Impact on population stratification

When CMR and echo LVEF were classified into three categories (<40, 40-50 and >50% EF), Cohen’s kappa agreement to CMR EF labels was similar between manual and DL1 contouring (0.65±0.11 vs 0.61±0.14) compared to 0.48±0.11 for DL2 and 0.29±0.11 for DL3. When confronting echo vs. CMR EF labels for A_1_’s frames, manual contouring misclassified 22 patients, and DL contouring misclassified 16 patients. However, for A_2_ and B_1_, the opposite situation was observed: there were 33 misclassified subjects using manual contouring for A_2_ vs. 27 using DL1 (35 vs. 19 for B_1_).

## Discussion

The principal findings of our study can be summarized as follows.

First, our DL algorithm (DL1) trained echo contouring performed well in an unrelated population of patients despite a balanced dataset in terms of image quality. It generalized well to data obtained in another centre (B) representing 80% of cases.

Second, DL1 reduced interobserver agreement relative to manual contouring for LV-EF, EDV and ESV, and in particular between junior and senior observers. This was confirmed for the other two compared DL algorithms, tending to suggest that DL can be instrumental in increasing the interobserver reproducibility of 2D echo-based EF values, if taken as an initial contour before potential manual edits, when required in more challenging cases.

Third, this study demonstrated that DL1 compared favourably to manual contouring in terms of EF accuracy when taking CMR as reference (Fig 6). EF bias was brought to almost zero when segmenting the same image frames as the 3 observers with DL1, which reduced the bias of one junior and one senior observer. Yet biases in LV volumes remained present and DL1 did not correct the underestimation of 2D echo-based volumes, compared to CMR. Such underestimation is believed to result in part from foreshortening of acquired 2D, this bias is known to be reduced by 3D echo. It may also result from differences in detection of trabeculated myocardium. Accordingly, DL algorithms trained with both echo and CMR data might allow learning some of this systematic bias. Adding more pathological groups to the training database could potentially improve EF biases for different disease groups.

When comparing our DL results with previously trained network, we found (Fig 7) better agreement with CMR EF and reduced bias than DL2 and DL3. However, DL2 appeared as a clear contender and showed excellent inter-observer agreement and a good correlation with CMR. It can be argued the comparison done in this paper is unfair, as DL1 was trained on data from clinical centre A, all performed on Philips echocardiographic devices, with manual contouring from observer A1. DL2 was trained on ground truths segmentations from other observers and on GE data. Therefore, the comparison presented here should be taken as a direct test of generatability of an echocardiographic DL segmentation algorithm (DL2) without applying any transfer learning to another constructor and possibly with other contouring conventions. Our results illustrate the need to learn models that generalize well across vendors and clinical centres, possibly through federated learning. DL3 was applied similarly without any and adaptation and performed poorly on our data. As for DL2, this probably reflects discrepancies between the training data of DL1 and DL3 and calls for further adaptation of the DL3 network to DL1 training data that are beyond the scope of this paper.

Finally, using CMR-based EF reference values, we could evaluate the potential impact that an echo- and DL-based EF computation would have on the stratification of a Heart Failure population. We found a similar (A_1_) or improved (A_2_, B_1_) agreement between echo measurements vs. CMR using the DL algorithm over manual contouring. This preliminary finding should be confirmed on a population of HF patients with preserved and reduced EF to determine whether or not the added value of DL vs. manual contouring is confirmed.

Several previous studies did report agreement and correlation values of automated and manual EF values. The AutoEF algorithm was evaluated in large studies (>200 subjects (21)) but the comparison to CMR could only be performed for a subset (∼20) of the population. In (22), the AutoEF results were edited when deemed necessary by both senior and novice observers, which represents a potential bias when comparing manual and automated contouring solutions. Other commercial algorithms (23) were also assessed against manual contouring but often without involving another modality as reference. As DL-based segmentation solutions are emerging in echocardiography (13,24), they need to be benchmarked not only for accuracy against manual observers but also against other imaging modalities, and more specifically against CMR at it stands as a gold standard modality for LV EF assessment like CMR.

Most of EF validation studies that took CMR as reference were comparing 2D echo to 3D echo, and demonstrated a higher accuracy on EDV and ESV measurements (24), as well as lower intra- and interobservers variability (25) and higher performance for some pathologies such as HCM (26). Nonetheless, the spatiotemporal resolution of 3D echo, which is inherently lower than 2D imaging, can be challenging with larger chambers. In addition, 3D echo remains a premium imaging modality, not as widespread as 2D echo. Improving the echocardiographic workflow involves automating time-consuming tasks for 2D echo images as well as 3D echo. However, the processing of 2D echo is still mostly manual, unlike 3D echo, for which advanced model-based (editable) segmentation algorithms are available (25,26). This situation called for a thorough evaluation of a modern automated segmentation on 2D echo, validating it with another 3D reference modality.

By contributing an open validation dataset, together with the bi-plane Simpson code, this paper contributes a reproducible evaluation framework, against which other DL methods can be benchmarked.

### Clinical implications

We argue that the framework described here could help exploit the full potential of deep learning for echocardiographic applications

- simplifying LVEF and volume calculations to allow for multi-cycle or real-time assessment.

- Improved longitudinal follow-up of chronic patients due to good overall agreement with CMR and reduced inter-observer variability.

- Improved management strategies due to the accuracy of the LVEF category classification.

### Study Limitations

In this paper, the observers, not to interfere with clinical practice, were free to choose the cycles and frames on which they quantified EDV and ESV volumes. Therefore, we could not compute local contour differences between observers. Such a local analysis could have revealed regions of higher variability or systematic interobserver differences. Similarly, we could not study if the DL segmentation represents a good consensus by comparing its contour to the observers’ contours. A further automatization could include a separate pre-processing DL network automatically selecting the ES and ED frames. This was left as future work and likely requires a separate evaluation.

We limited our comparisons to EDV, ESV and EF values, as they appeared as a priority, being clinical indices used routinely. Yet this approach probably better reflected clinical practice, where there is also intersubject variability in selection of frames. The clinical centres compared in this study have similar protocols in terms of echocardiography and used the same equipment. The algorithm might behave less accurately on other echocardiographic systems or image acquisition protocols. Extending the analysis of this paper to other clinical centres could further span differences across countries in terms of conventions for defining the endocardial contour, in terms of expertise (e.g. junior sonographer vs senior cardiologist), or in terms of time constraints for the echocardiographic exam. Also, we did not cover in this study reproducibility issues stemming from the acquisition (e.g. probe orientation) that can induce foreshortening. Finally, although CMR is widely accepted as reference modality for the validation of echo-based measurements, measurements performed on short axis slices only could underestimate the long axis contribution of LV motion (27).

## Conclusions

In this paper, we compared manual and DL automated contouring from 2D echocardiographic images with respect to CMR, taking the latter as reference for the computation of EF, ESV and EDV values. We demonstrated the value of a DL-based automated contouring of AP2 and AP4 images to reduce and homogenize the biases in EF with respect to CMR. This study also confirmed important biases in EDV and ESV 2D echo-based values, for automated and manual contouring, that nonetheless get compensated when computing EF, reaching a practically null bias between CMR and echo-based EF values.

## Data Availability

All DICOM files of AP2, AP4 images will be made available upon acceptance on a public web page. An excel file containing all quantification from MR will also be made available.

## List of Abbreviations and Acronyms

AP4: apical 2-chamber
AP2: apical 4-chamber
CCC: Lin’s concordance correlation coefficient
CMP: cardiomyopathy
DL: deep learning
EF: ejection fraction
ED[V]: end-diastolic [volume]
ES[V]: end-systolic [volume]
LV: left ventricle
VOL: healthy volunteer

## Funding

This research received no specific grant from any funding agency in the public, commercial, or not-for-profit sectors.

## Supporting information

**S1 Fig. Overlay of DL segmentations (yellow mask) and manual contouring (red dotted contour) for 6 outliers**. Frame and manual segmentation performed by observer A_1_ (top row), A_2_ (middle row) and B_1_ (bottom row). Outliers involved subjects with dilated non-ischaemic cardiomyopathy (NON-ISCH), mitral regurgitation or aortic stenosis (VALV) and hypertrophic cardiomyopathy (HCM).

**S2 Fig. Lin’s concordance correlation plots between CMR and 2D echo for the DL2 and DL3 algorithms applied to the frames of each observer** (A_1_ [senior], A_2_ [junior] and B_1_ [senior]) for LV EDV and LV ESV.

**S1 Table. Agreement between observers in echo for ESV, EDV and EF with manual and DL contouring using Lin’s concordance correlation coefficient**

## Notes

### Competing Interest Statement

The authors have declared no competing interest.

### Funding Statement

The authors received no specific funding for this work. Alexandre Popoff (AP), Mathieu De Craene (MDC), Guillaume Pizaine GP) and Pascal Allain (PA) are employed by Philips Medical Systems. Hélène Langet (HL), Romane Gauriau (RG) and Paulo Piro (PP) were employed by Philips Medical Systems at the time of the study. Centre Hospitalier Universitaire Caen Normandie and Cliniques Universitaires Saint-Luc have a master research agreement with Philips France Commercial.

### Author Declarations

The studies were approved by the IRB in charge (either Comité Ethique Hospital- Facultaire de l’Université Catholique de Louvain, Brussels, Belgium or Comité de Protection des Personnes Nord-Ouest III, Caen, France). The retrospective use of these data satisfies the European Union (EU) General Data Protection Regulation (GDPR) requirements.

